# Mixed-Frequency Regression Model for Short-Term Environmental Exposure–Response Modelling: A Simulation Study

**DOI:** 10.64898/2026.06.24.26356336

**Authors:** Nidhi Shukla, Hadiqa Tahir, Simon Smart, Suzanne E. Bartington, Anna L. Hansell, Tim CD Lucas

## Abstract

**Background:** Extreme environmental events, such as extreme temperatures and air pollution, have become a global concern due to their detrimental effects on human health. Short-term peak exposure episodes, despite lasting only a few hours, are crucial for exposure-response modelling. The use of time-aggregated exposure data often overlooks the impact of peak exposures on human health. However, studies employing high-temporal resolution exposure data are rare due to the limited availability of high-temporal resolution health outcomes across various scenarios. Therefore, to address the limitations associated with exposure-response modelling using aggregated exposure data, we have developed a model referred to as the mixed-frequency distributed lag non-linear model (mf-DLNM).

**Methods:** In this work, a simulation study was conducted to further validate the mf-DLNM for hourly-daily mixed-frequency data, using data on hourly temperature and daily respiratory mortality for the West Midlands, UK. Given that the focus was on extreme exposures, Relative Risks (RR) at the 5th and 95th temperature quantiles were considered as the estimands of interest. Model performance was evaluated based on the bias, empirical standard error (EmpSE), and coverage of these estimands. Additionally, the model was assessed across various scenarios, considering data size (1, 3, 5, and 11 years with a 24-hour lag), lag length (12 and 24 hours with 11 years), seasonal variation (summer months with 11 years and 24-hour lag) and distribution (Poisson and negative binomial).

**Results:** The mf-DLNM effectively captured the true parameters of the model. The model, fitted to 11 years of simulated data, a 24-hour lag and a Poisson distribution, observed a bias of 0.011 (0.0009) and 0.011 (0.001) for the RR at the 5th and 95th temperature quantiles, respectively, with Monte Carlo SEs (MCSEs) in parentheses. Furthermore, the model exhibited coverage of 0.94 and 0.93 for RR at the 5th and 95th temperature quantiles, respectively. In addition, the mf-DLNM with hourly and daily data demonstrated satisfactory performance across all scenarios except for the RR at 95^th^ temperature quantiles in the seasonal analysis.

**Conclusions:** Researchers are encouraged to adopt mf-DLNM in instances where high-temporal resolution exposure data are available alongside low-resolution health data. It serves as an alternative to traditional approaches that aggregate high-frequency exposure data. By preserving the temporal information of environmental exposures, mf-DLNM enables a more precise assessment of exposure-response relationships, thereby improving the accuracy and reliability of health risk estimates. This approach offers a promising opportunity for informed decision-making and the development of effective interventions for vulnerable populations and healthcare facilities to address short-term environmental episodes.

## 1. Introduction

The primary motivation for exposure-response modelling studies is to safeguard human health from adverse environmental exposures, and the selected time scale i.e., short or long-term critically influences the interpretation of these studies.^1^ Short-term exposure modelling studies typically span from days to a few weeks, whereas long-term studies extend over weeks to years.^2,3^ However, to accurately assess acute health risks associated with short-term episodes, it is essential to examine the hourly variability of environmental exposures.

Globally, several studies have documented peak environmental episodes. For instance, Beijing experienced an extreme air pollution episode in 2013, with an hourly PM_2.5_ concentration exceeding 600 μg/m^3^.^4^ Similarly, in 2017 Delhi recorded an hourly PM_2.5_ concentration high as 770 μg/m^3^.^5^ In the UK, a severe air pollution episode in 2014 led to recorded hourly PM_10_ values of up to 100 μg/m^3^ in 2014.^6^ Extreme temperature events have also been reported, such as heatwave in China, where maximum temperature up to 41.1 °C, which was 6.1 °C higher than the country’s heatwave threshold.^7^ In addition, metropolitan cities in India have also experienced significant heatwave episodes.^8^ Given the global occurrence of short-term spikes in air pollutants and temperature changes, exploring their impact on human health has become crucial. Furthermore, the short-term exposure to extreme temperature and air pollution are individually associated with increased risk of mortality, while their co-exposure have larger effects.^9^

A few studies have demonstrated the association between hourly variations in environmental exposure and subsequent health impacts. The exposure to PM_2.5_ may increase the risk of cardiorespiratory hospital admission within hours of exposure.^10^ Furthermore, hourly peak PM_2.5_ was reported to be associated with all-cause mortality as well as same-day personal exposure to PM fractions are also associated with asthma-related symptoms^11,12^. Furthermore, a study observed that walking for 2-hours in polluted environments could lead to reductions in the forced expiratory volume in 1 second (FEV1).^13^ Similarly, another study found that a 25 μg/m³ increase in PM_2.5_ over a two-hour period was strongly associated with the onset of acute myocardial infarction.^14^

Temperature variability also significantly impacts human health.^15^ A study analysing data from 45 US metropolitan areas (1987-2000) found that a unit increase in hourly temperature variability was associated with 0.53% (95% CI: 0.13-0.94%) increase in respiratory mortality risk.^16^ A study reported similar findings in Australia, where a unit increase in hourly temperature variability raised mortality risk by 0.51% (95% CI: 0.33–0.69%), contributing to 1.67% of nationwide deaths annually.^17^ Additionally, both high and low diurnal temperature ranges (DTR) have been linked to increased asthma hospitalizations, with high DTR significantly elevating asthma hospitalization risk in adults.^18^

Despite these findings, studies incorporating complete temporal patterns remain rare due to the unavailability of hourly health outcome data. Metrics such as DTR, which summarize 24-hour temperature variation using the difference between daily maximum and minimum temperatures, oversimplify hourly changes. This as above ignores important hourly temperature variation and may not provide complete association. Furthermore, many studies on hourly temperature variability (HTV) rely on standard deviation of hourly temperature rather than direct measurements of hourly temperature variations.

To address this limitation, there is a need for statistical regression models capable of handling mixed-frequency data in environmental epidemiology. The concept of mixed-frequency regression modelling has been initially applied in the field of econometrics ^19^, where it is referred to as MIDAS (Mixed Data Sampling). MIDAS models are similar to Distributed Lag Models (DLMs).^20^ Distributed Lag Models (DLMs) are statistical approaches used to assess the delayed or lagged effect of an exposure on an outcome. The key distinction between MIDAS and DLM, is that DLM is same frequency regression model while MIDAS can handle mixed-frequency data. An extension of the DLM has been developed, known as the Distributed Lag Non-Linear Model (DLNM).^21^ DLNMs represent a more advanced and suitable approach for applications in environmental epidemiology, as they allow for the estimation of non-linear relationships in both the exposure–response and lag–response dimensions. Some recent literature has used the DLNM for mixed-frequency regression involving daily and weekly mixed frequency data.^22,23^

This work proposes utilizing the DLNM framework for mixed-frequency modelling as an alternative method to investigate the association between high-frequency hourly exposure data and low-frequency health daily outcomes. We evaluate the performance of the mixed frequency (mf)-DLNM for hourly-daily data exposure response modelling, by applying the model to hourly exposure (temperature) and daily response (respiratory mortality) data. In the absence of actual hourly respiratory mortality data, a direct comparison between observed and predicted values was not possible. Instead, we conducted a simulation study to validate the mf-DLNM for hourly-daily mixed-frequency modelling. To evaluate this approach, we utilized ERA-5 land temperature data from the European Centre for Medium-Range Weather Forecasts (ECMWF) Copernicus and respiratory mortality data from the Office of National Statistics (ONS) for the West Midlands, UK.

## 2. Method

### 2.1 Overview

This section outlines the methodological framework adopted in this study. First, we present the framework of the mf-DLNM. Subsequently, the baseline mortality risk, assumed as true risk, was obtained by fitting the mf-DLNM to hourly temperature and daily respiratory mortality data from January 01, 2010 to December 31, 2020 for the West Midlands region of the UK. This fitted model is used throughout the subsequent simulation study as the ground truth. We then assessed the proposed mixed-frequency hourly-daily framework of the mf-DLNM using a simulation study. Model performance was evaluated under different exposure-response scenarios. Given the importance of peak environmental exposure, the RR at the 5th and 95th quantiles of the temperature distribution were used as estimands to assess performance under extreme exposure conditions.

### 2.2 Mixed frequency mf-DLNM

This model was developed using the concept of DLNM and MIDAS. It is similar to DLNM with a major distinction in the frequency of the regressor and the outcome. A typical mf-DLNM is expressed as follows;

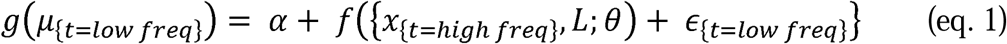

Here, y_t_ is the outcome at time t and is assumed to be from a distribution in the exponential family where µ_t_ = E(y_t_). Further, g is a monotonic link function, and x_t_ is the regressor. Given that this is a mixed frequency model y_t_ and x_t_ are at different temporal frequencies, such as hourly and daily, respectively. f(x_t_, L; θ) defines the cross-basis matrix for exposure and lag, which is a product of the basis function of exposure and lag.

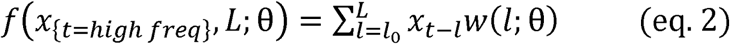

Therefore, the function b.w (x_t-l_, l; θ) combines an exposure-response function b(x) and a lag-response function w(l), together expressing a bi-dimensional basis function referred to as the cross-basis.^21^

### 2.3 Baseline Model

The baseline RR was computed using the mf-DLNM, fitted to hourly temperature and daily respiratory mortality data from January 01, 2010 to December 31, 2020. The cross-basis function with B-splines was applied to both the temperature and lag dimensions. For the temperature dimension, B-splines with equal knots were used, with 6 degrees of freedom (df). For the lag dimension, B-splines with equally spaced log-values were applied, with 5 df. The model was fitted with a maximum lag of 24. A generalized additive model (GAM) with Poisson regression was employed to estimate the temperature-mortality association. The long-term trend and the seasonality was controlled in the model using a smooth function (natural spline) of time with 7 df per year. Thus, these smooth functions control for the seasonality and between-year patterns. The specification of the time component varied across different scenarios.

The simulation was validated under multiple scenarios. Therefore, separate baseline models were run for each scenarios including variation in data sizes (1, 3, 5, and 11 years while holding the number of lags constant at 24), lag lengths (12 and 24 hours while keeping the sample size constant at 11 years), seasonal variation (summer months with 11 years of data and 24-hour lag) and distributions (Poisson and negative binomial).

### 2.4 Simulation Study

A simulation study was conducted to assess the performance of the mf-DLNM in capturing the relationship between hourly exposure and daily health outcome data. The study followed the ADEMP protocol as given in **Table 1**, defines the aims, data-generating mechanism (DGMs), estimands, methods, and performance measures of the study.^24^

**Table 1:** ADEMP structure of the simulation study.

| Steps | Decisions |
| --- | --- |
| Aim | Validate the mf-DLNM for hourly (exposure)-daily (response) mixed frequency regression |
| **Data Generating Mechanisms (DGMs)** | Simulated data were generated using original temperature data and parameters retrieved from the baseline model. The main model (referred to as **M1**) included 11 years of data, 24-hour lag, and Poisson distribution. Additionally, different DGMs (S1-S6) were used, including the following scenarios:<br>▪ Data size (1, 3, 5, and 11 years with a 24-hour lag) [**S1, S2, S3, M1**]<br>▪ Lag length (12 and 24 hours with 11 years) [**S4, M1**]<br>▪ Seasonal variation (summer months with 11 years and 24-hour lag) [**S5**]<br>▪ Distribution (Poisson and negative binomial) [**S6, M1**] |
| **Estimands** | RR at the 5th and 95th percentile of temperature |
| **Method** | mf-DLNM |
| **Performance measures** | Bias, empirical standard error (EmpSE), and Coverage |

#### 2.4.1. Aims

The primary aim of the simulation study was to validate the use of mf-DLNM for estimating the relationship between high temporal-resolution exposure and low temporal resolution outcome data (i.e., hourly exposure and daily response data). This approach holds significant potential for exposure–response modelling, as it enables the use of high-resolution hourly exposure data even when similar health data are unavailable. Ultimately, this framework enhances public health preparedness by improving the prediction of health impacts due to extreme environmental events.

#### 2.4.2. Data generation

Firstly, a baseline mf-DLNM model was fitted using observed hourly temperature and daily mortality data, as discussed in **Section 2.3**, across different scenarios outlined in **Table 1**. The model parameters from baseline model and true temperature data were used to generate data, assuming a Poisson distribution, via the rpois() function in R. The simulation study primarily focused on the model fitted using 11 years of hourly temperature and daily mortality data, with 24 hours lag, and a Poisson distribution, referred as main model (M1) in this work. These true parameters from the baseline model were used to generate 1000 sets of mortality data. Various scenarios were assessed to further strengthen the model validation through this simulation study. The baseline model was fitted for different scenarios including different data sizes (1, 3, 5, and 11 years, with a 24-hour lag and Poisson distribution), different lag lengths (12 and 24 hours, with 11 years of data and Poisson distribution), a single season (summer months over 11 years, with a 24-hour lag and Poisson distribution), and different distributions (11 years with a 24-hour lag and a negative binomial distribution). Similar to the main model, the model parameters from these individual baseline frameworks, along with respective temperature data were used to generate the response variable, through 1000 repetitions. Data generation was performed using the parameters from these models, following the Poisson distribution, except for scenario 6, where a negative binomial distribution was used.

#### 2.4.3. Estimands

The RR at the 5th and 95th temperature quantiles were selected as the estimands to evaluate the model performance. Studies assessing temperature-related health risks typically estimate effects at extreme temperatures, with cold effects at lower percentiles and heat effects at higher percentiles.^25^ Although the 1st and 99th percentiles capture most extreme events, they may not fully characterize the broader temperature-mortality relationship.^26^ Therefore, the 5th and 95th temperature quantiles were selected as estimands, representing a reasonable value for assessing both heat- and cold-related mortality risks.

#### 2.4.4 Methods

The mf-DLNM was implemented using a GAM with DLNM following a Poisson distribution. The model was fitted in the same way as equation (1), with hourly temperature data and daily mortality data. Different models were fitted for different scenarios, considering respective specifications.

#### 2.4.5 Performance measures

Performance metrics, including bias, empirical standard error (EmpSE), and coverage, along with their respective Monte Carlo standard errors (MCSEs), were evaluated. Furthermore, the study also examined the model standard error (ModSE) and respective MCSE. The performance evaluation was based on 1000 repetitions, ensuring sufficiently low MCSEs.

This detailed simulation process is summarised in the **Figure 1**. To assess the sensitivity to assumptions about the shape of the lag structure and the length of the lag interval, the simulated model was run for varying numbers of knots both in the lag and exposure dimension. Given the mixed-frequency nature of the model, sensitivity was specifically evaluated using different numbers of knots in the exposure dimension.

**Figure 1:**
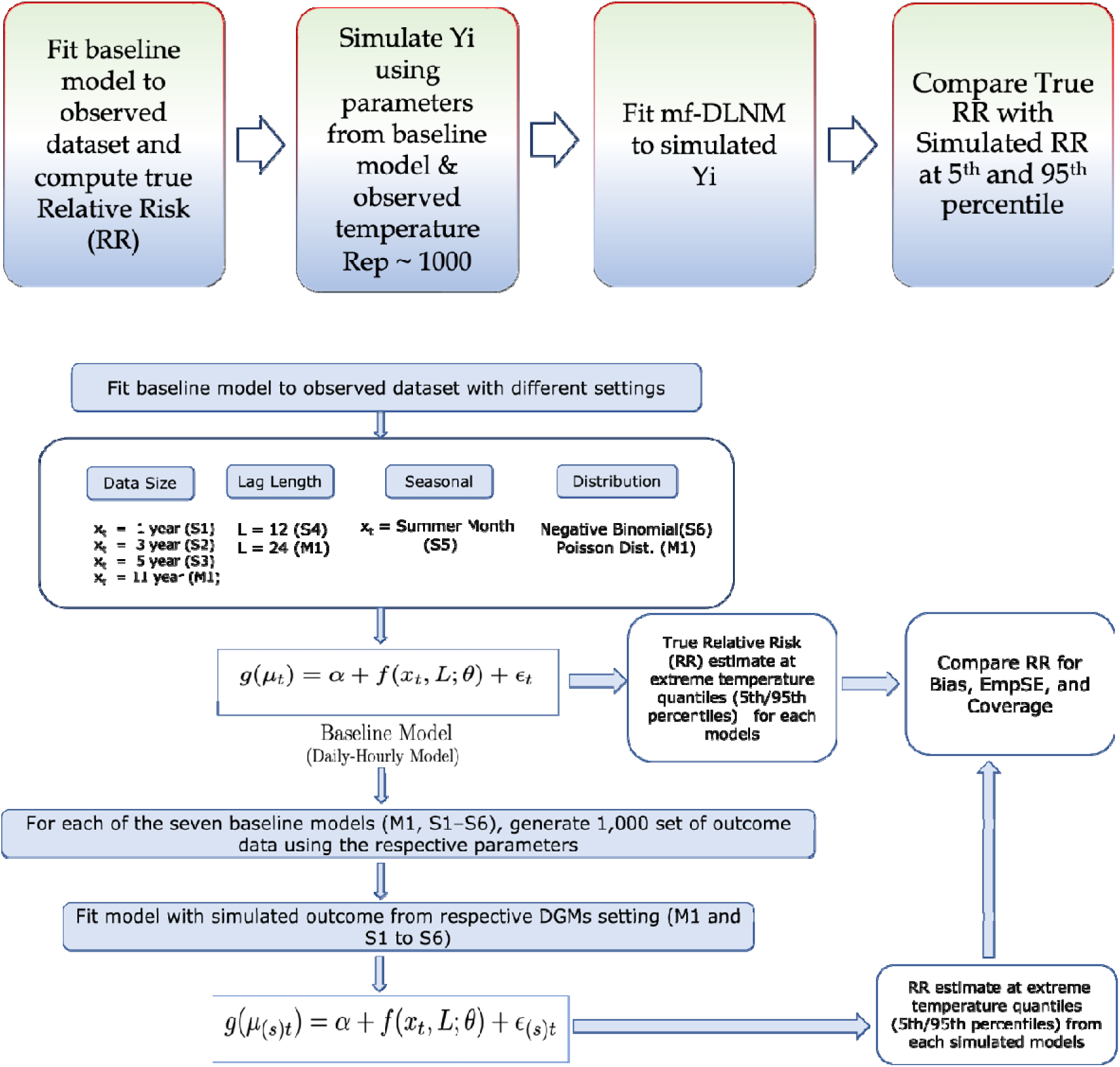
Flow chart to demonstrate the method for simulation study.

## 3. Results

### 3.1. Descriptive data

The total respiratory mortality count for the West Midlands from study period was 78,957. The mean, mean minimum, and mean maximum temperatures during this period were 8, -10, and 30, respectively. **Figure 2** illustrates the distribution of hourly temperature and respiratory mortality, showing that the mortality count had a moderately right-skewed distribution.

**Figure 2.**
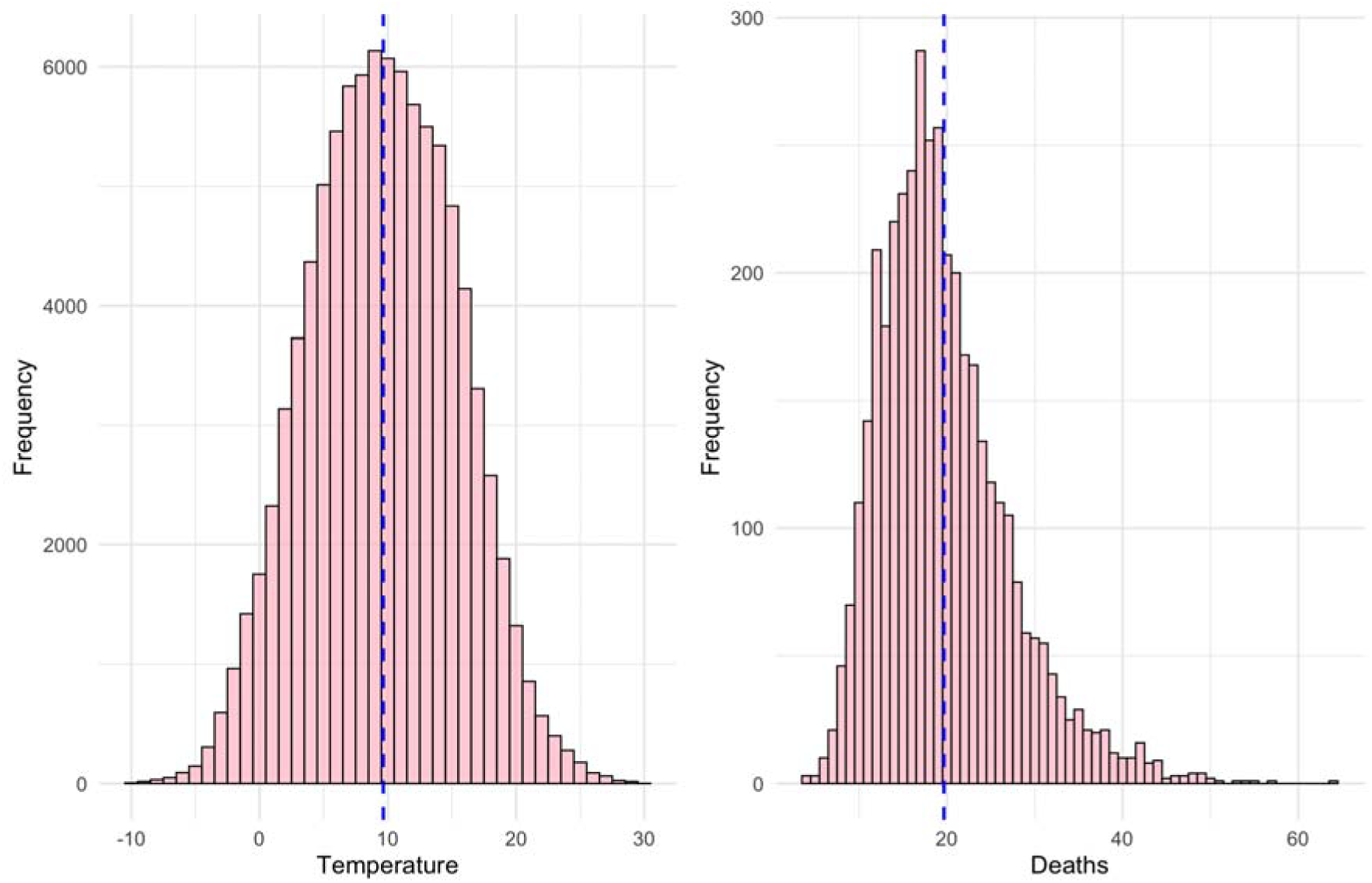
Distribution of hourly temperature and respiratory mortality counts for West Midlands, UK from 2010 to 2020.

### 3.2 Estimates of baseline model

This section presents the relationship between hourly temperature and daily mortality using the baseline model. The analysis indicates that temperature does not have immediate impact on mortality, while extreme heat effects were observed with a 3-hour lag. Cold temperatures did not exhibit an immediate impact on mortality within the 24 hour lag period. The overall cumulative temperature effect, which accounts for contributions over the 24-hour lag is illustrated in **Figure 3**. The overall RR for the 95th and 99.9th temperature quantiles were 1.07 (95% CI: 0.99–1.16) and 1.35 (95% CI: 0.95–1.91), respectively. Furthermore, the overall cumulative risk for a 5-unit increase in temperature was 1.10 (95% CI: 0.99–1.23) over a 24-hour lag. It is important to emphasize that this risk assessment was conducted solely for methodological evaluation and not intended to inform policy development. Rather, the primary objective was to assess the performance of the mf-DLNM in the context of mixed-frequency modelling within environmental epidemiology.

**Figure 3.**
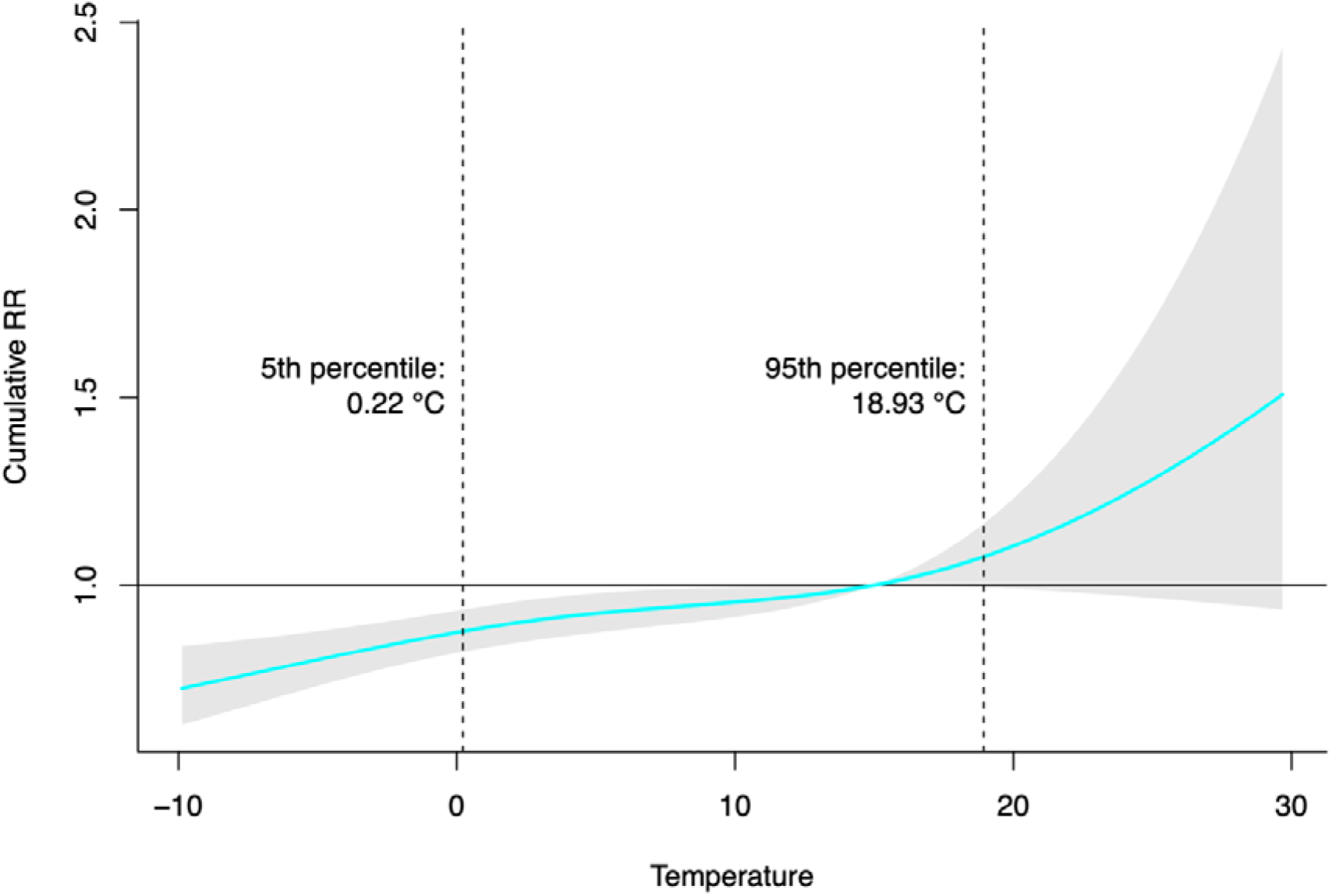
The cumulative exposure-response curves for the associations between temperature and respiratory mortality over a lag of 0–24 hours using a reference temperature of 15°C. The red line shows the cumulative relative risk (RR) of respiratory mortality, and the grey areas are the 95% confidence intervals.

### 3.3 Results for model performance

This section presents the model performance results by assessing the comparison of model parameters between the baseline models and the models fitted to simulated data. The models fitted to simulated data referred as simulated model for convenience of reporting. Furthermore, it evaluates the model’s performance in estimating the RR of respiratory mortality associated with the 5th and 95th temperature quantiles, primarily using three performance metrics: bias, empirical standard error (EmpSE), and coverage.

#### 3.3.1 Comparison of model parameters

The estimated parameters from the models fitted to simulated data (referred as simulated model) and the baseline model were compared using Pearson correlation coefficients. **Figure 4** illustrates the correlation between parameters from 20 simulated models and the baseline model, demonstrating that the simulated model accurately captures the characteristics of the baseline model. The Pearson correlation coefficient (R) was ≥ 0.96, indicating that the simulated model effectively replicates the baseline model parameters with high accuracy.

**Figure 4.**
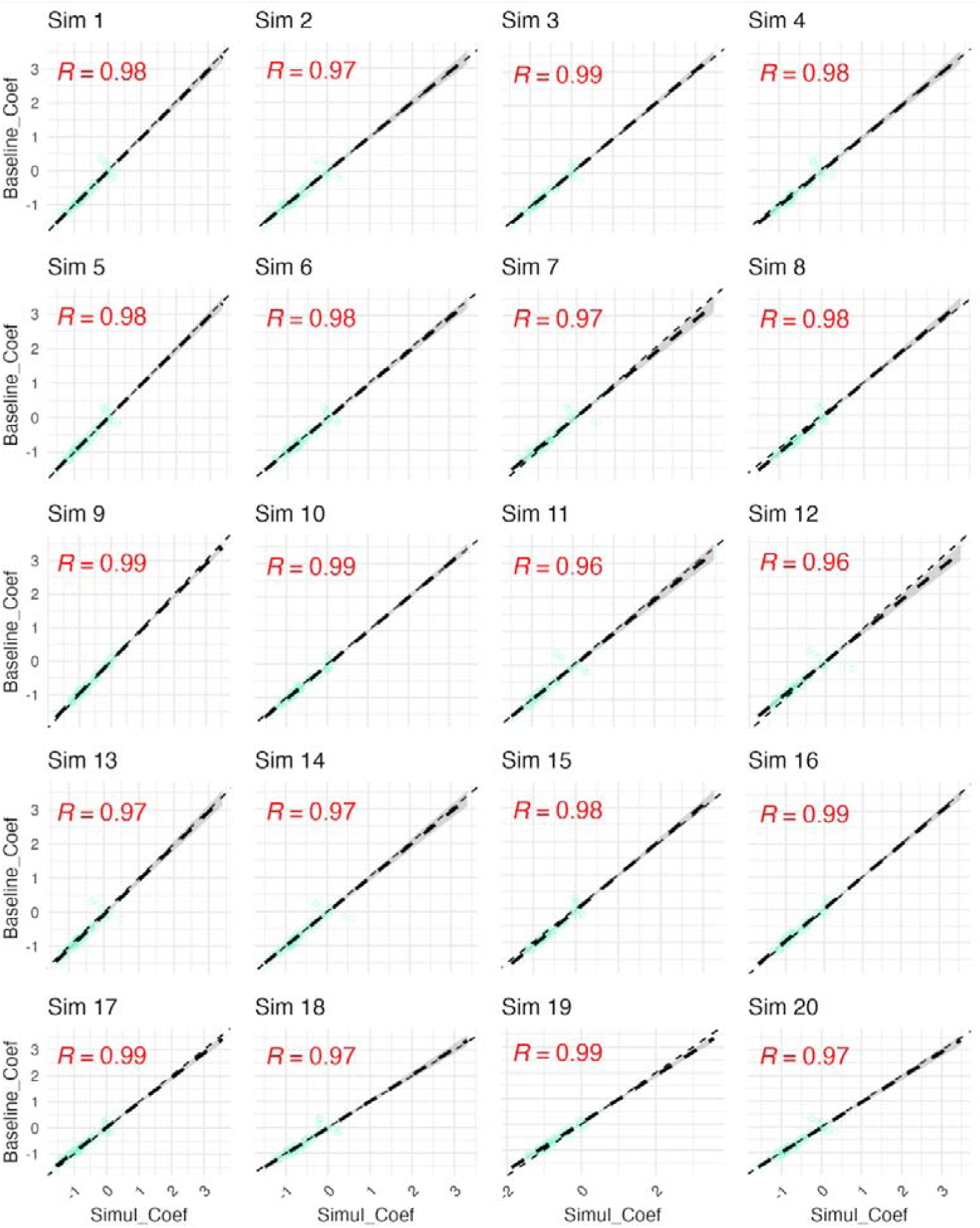
Model performance for nsim=20 parameters from baseline and simulated model.

#### 3.3.2 Estimands performance

The model was fitted to 1000 sets of mortality counts generated using the DGMs and observed hourly temperature data to estimate the RR of respiratory mortality at the 5th and 95th temperature quantiles. Model performance was compared to that of the main baseline model, which was fitted using 11 years of temperature and mortality data with a 24-hour lag, assuming a Poisson distribution. The simulated model had a specification similar to that of the baseline model. **Figure 5** shows the deviation of simulated RR from the true RR.

**Figure 5.**
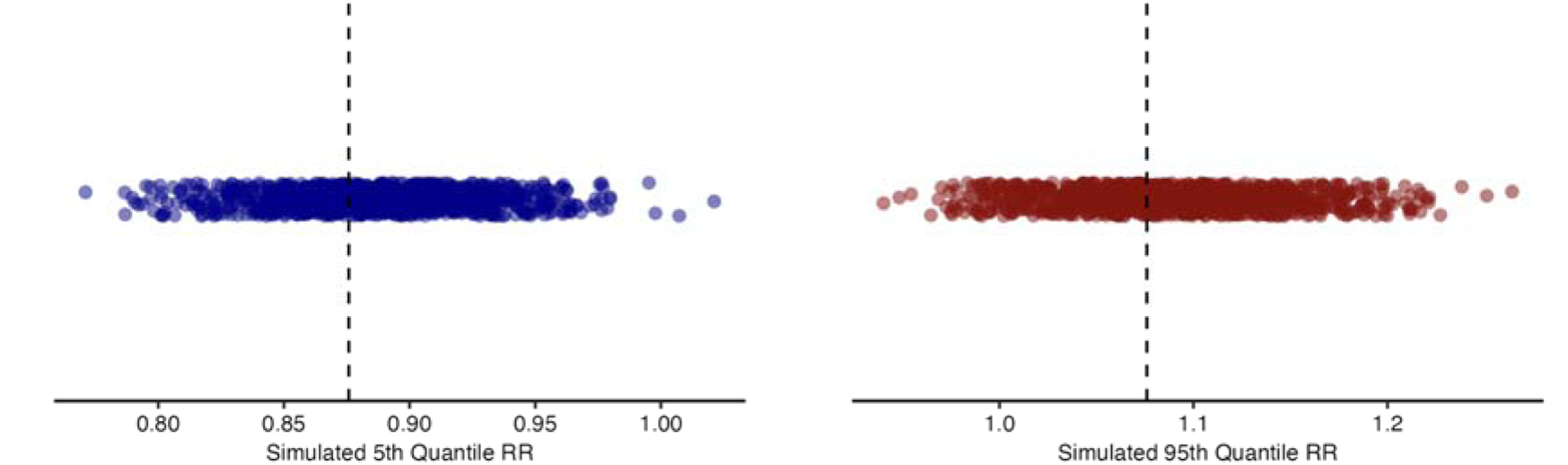
Plot of the 1000 estimates of Relative Risk (RR) at 5th (left panels) and 95th (right panels) temperature quantiles estimated from the 1000 sets of respiratory mortality data generated using data-generating mechanism M1. The dashed black vertical line represent the true RR for respective quantiles from the baseline model.

The bias for the RR at the 5th and 95th temperature quantiles was observed to be 0.011 (0.0009) and 0.011 (0.001), respectively, with MCSEs in parentheses. This indicates a low positive bias and small MCSEs. The EmpSE for the 5th and 95th temperature quantiles of RR was observed to be 0.028 (0.0006) and 0.044 (0.001), respectively, with MCSE in parentheses. The low estimated values of EmpSE suggest that the results are consistent across different simulations, while the low MCSE indicate minimal uncertainty. Furthermore, the results showed a low deviation between EmpSE and ModSE for the estimands of both quantiles. These performance measures are illustrated in **Figure 6**, and a summary is provided in **Table 2**.

**Figure 6.**
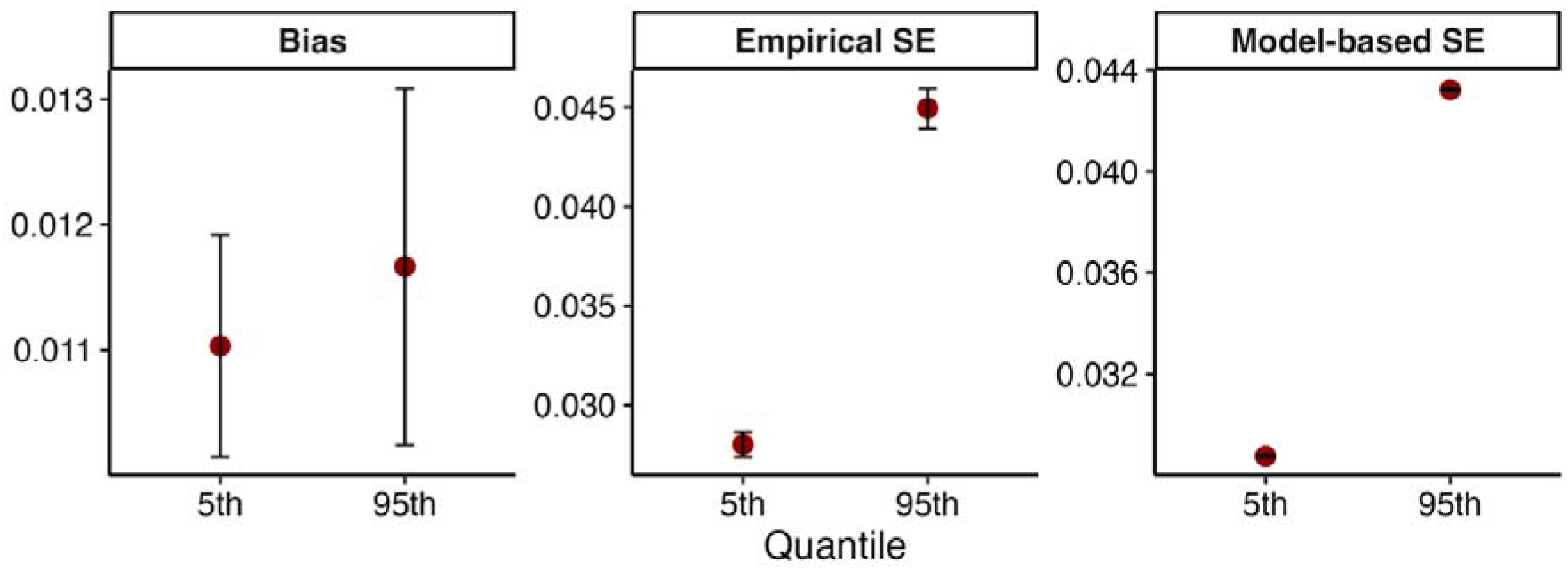
Plot of performance for measures of interest (Monte Carlo 95% confidence intervals as error bars).

**Table 2:.** Summary of method performance measures (Monte Carlo SEs in parentheses) for the main model with 11 years of data, 24-hour lag and Poisson distribution.

| Estimands | performance measures | Estimate (MC SEs) |
| --- | --- | --- |
| 5<sup>th</sup> Quantile | Bias | 0.011 (<0.001) |
|  | EmpSE | 0.028 (<0.001) |
|  | ModSE | 0.029 (<0.001) |
|  | Coverage | 0.945 (0.007) |
| 95<sup>th</sup> Quantile | Bias | 0.011 (0.001) |
|  | EmpSE | 0.045 (0.001) |
|  | ModSE | 0.043 (<0.001) |
|  | Coverage | 0.933 (0.007) |

The coverage is defined as the probability that a confidence interval contains the true estimate. It was computed as a binary function (0 for no coverage, 1 for coverage) for simulated risks at the temperature quantiles. **Figure 7** illustrates the coverage of the RR at the temperature quantiles for 1000 simulated models, along with the mean coverage and MCSEs. The donut charts in the figure show that coverage was equal to 1 in most of the 1000 repetitions for both quantiles. The mean coverage was 0.94 and 0.93 for the RR at the 5th and 95th temperature quantiles, respectively, with an MCSE of 0.007. The low MCSE indicates that no additional repetitions were necessary.

**Figure 7.**
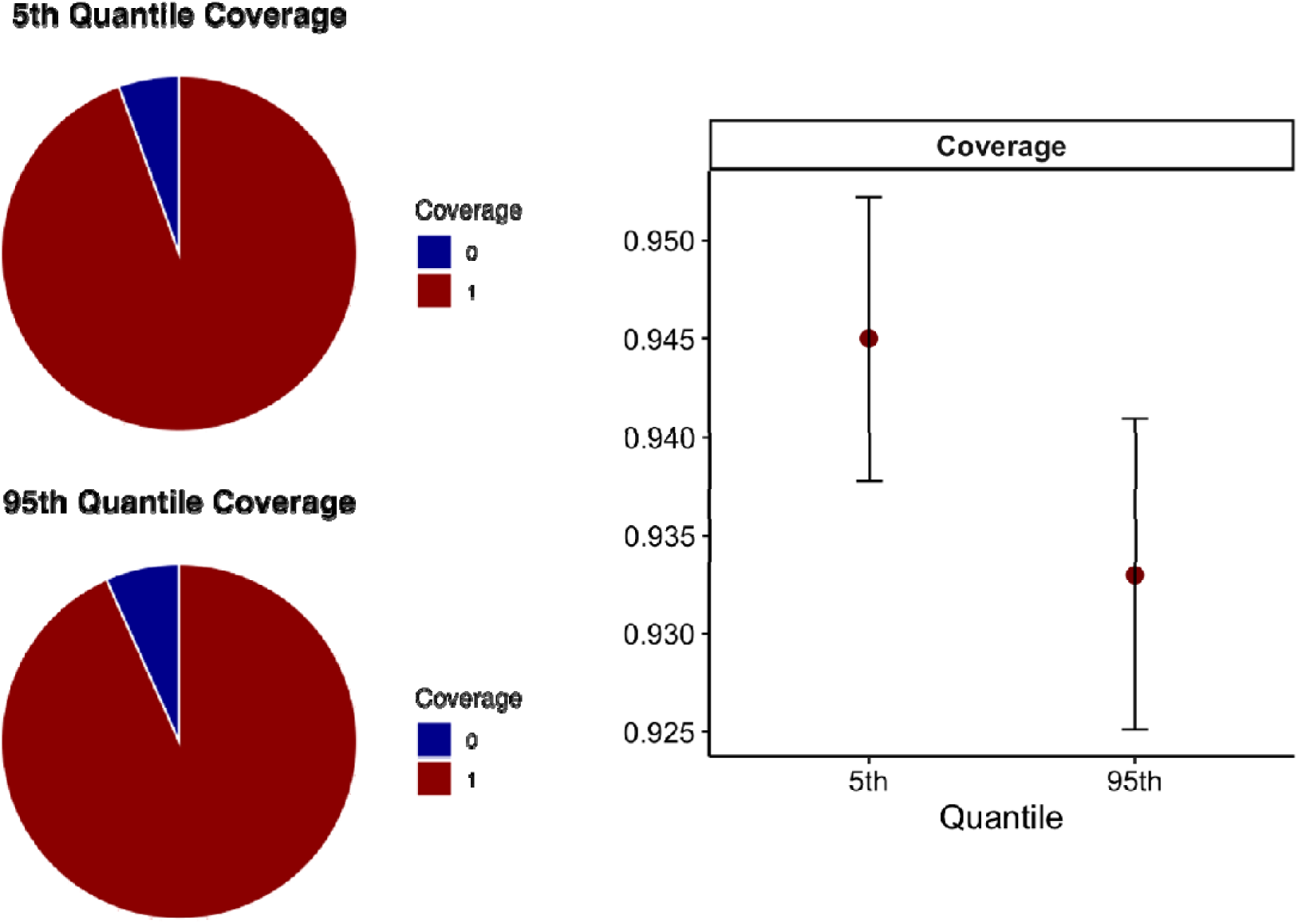
The left panel shows the coverage for 1000 sets of estimands, the Relative Risk (RR) at the 5th and 95th temperature quantiles, based on simulated model run with respective of respiratory mortality data generated using data-generating mechanisms. The right panel shows the average coverage for these estimands.

#### 3.3.3. Estimands performance for different scenarios

This section presents the results for various DGMs under different scenarios, with the corresponding performance measures depicted in **Figure 8**. When the mf-DNLM was applied using 1, 3, and 5 years of temporal data (Scenarios 1 to 3 in Table 1), the results indicated that as data size decreased, model deviation increased. Specifically, for the main model (M1), which was fitted using 11 years of data, the bias for the RR at both 5th and 95th temperature quantiles was 0.01. In contrast, in Scenario 1 (S1), which utilized only one year of data, the bias increased to 0.036 and 0.039 for estimating the respiratory mortality RR at the 5th and 95th temperature quantiles, respectively. The lowest bias was consistently observed in the main model with 11 years of data, compared to all data-size scenarios (S1-S3). However, no discernible pattern in coverage was observed across scenarios S1 to S3, with the main model (M1) demonstrating comparable coverage for the estimands at both quantiles. Scenario 4 (S4) investigated model performance as a function of lag length. The model exhibited biases of 0.015 (0.001) and -0.002 (0.001), and coverage values of 0.92 (0.008) and 0.93 (0.007) for the RR at the 5th and 95th temperature quantiles, respectively. These findings suggest that the model adequately captures the baseline model, even with a shortened lag length. Scenario 5 (S5), where the model was constrained to a single summer season, the model performance, particularly for the RR at the 95th temperature quantile, was found to be suboptimal compared to other scenarios, with higher bias of 0.138 (0.007) and an EmpSE of 0.20 (0.004). Furthermore, Scenario 6 (S6), assessed the effect of overdispersion by simulating data from a negative binomial distribution, using a theta value of 195 as derived from the baseline model. The estimated bias was 0.011 (0.0009) and 0.011 (0.001) for the RR at the 5th and 95th temperature quantiles, respectively. Coverage was 0.934 (0.007) and 0.945 (0.007) for the corresponding quantiles. These results indicate that model performance under Scenario 6 is comparable to that of the main model, which assumes a Poisson distributed outcome.

**Figure 8.**
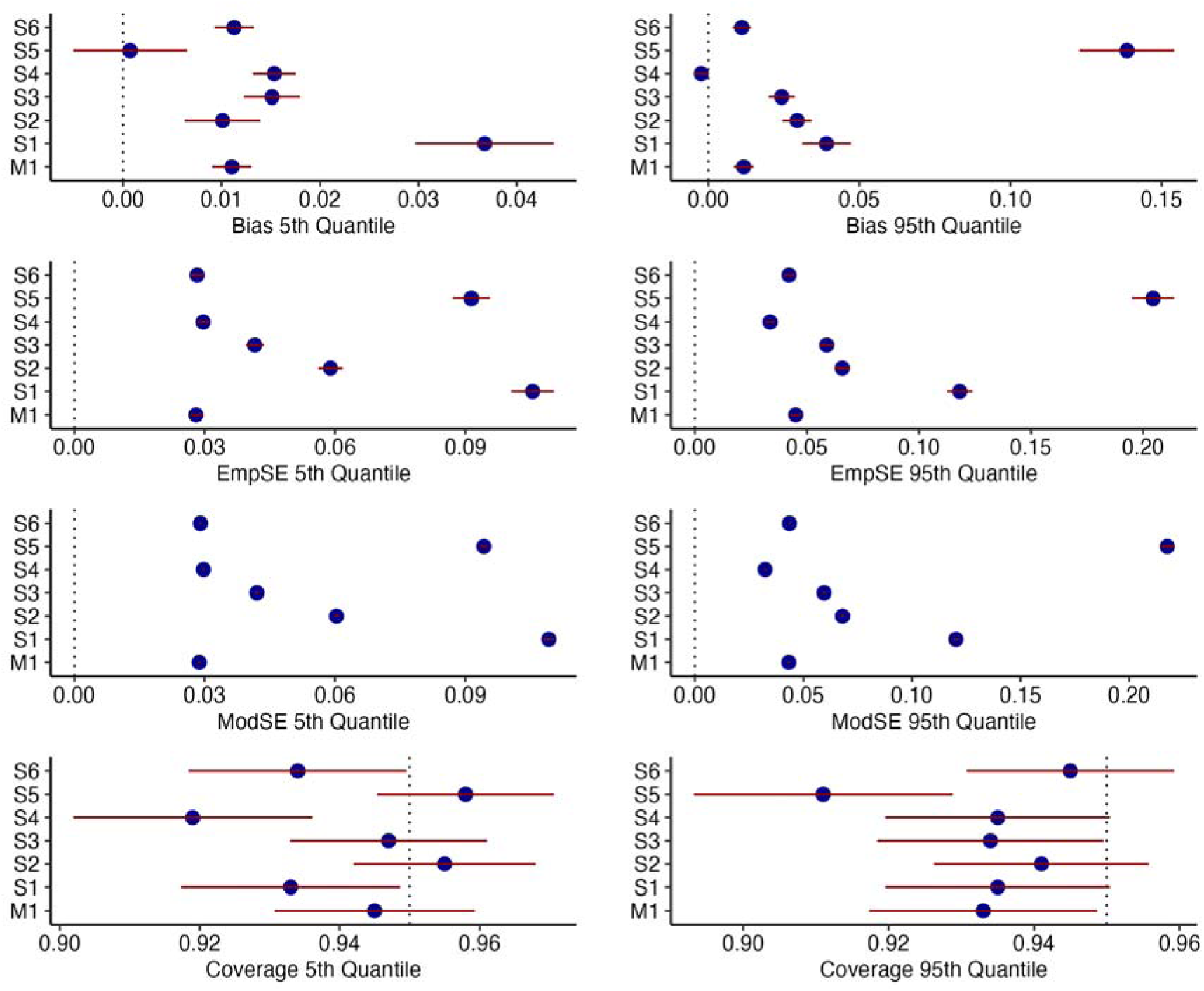
Performance measures and Monte Carlo 95% confidence intervals, with error bars, for the main model (M1) and scenarios assessment (S1-S6) with different DGM. For information on DGM refer **Table 1**.

### 3.4 Sensitivity Analysis

A series of sensitivity analyses (A1-A3) were conducted to evaluate the impact of alternative model specifications on performance. Specifically, modifications were made to the number of knots used in the dimension of predictor and lag dimension to assess the model’s robustness. The number of knots for the lag dimension was changed from 2 in the baseline model to 4 in the simulated model (A1), while the number of knots for the exposure dimension was increased from 2 in the baseline model to 3 (A2) and 4 (A3) in the simulated model.

The model performed well across all performance measures despite the change in the knot configurations, indicating its adequacy, as shown in **Figure 9**. The main model demonstrated that, across the three sensitivity analyses, the bias for the RR at the 5th temperature quantile ranged from 0.005 to 0.012, while the bias for the RR at the 95th temperature quantile ranged from 0.009 to 0.013. Coverage ranged from 0.94 to 0.95 for the RR at the 5th temperature quantile, and from 0.93 to 0.96 for the 95th quantile.

**Figure 9.**
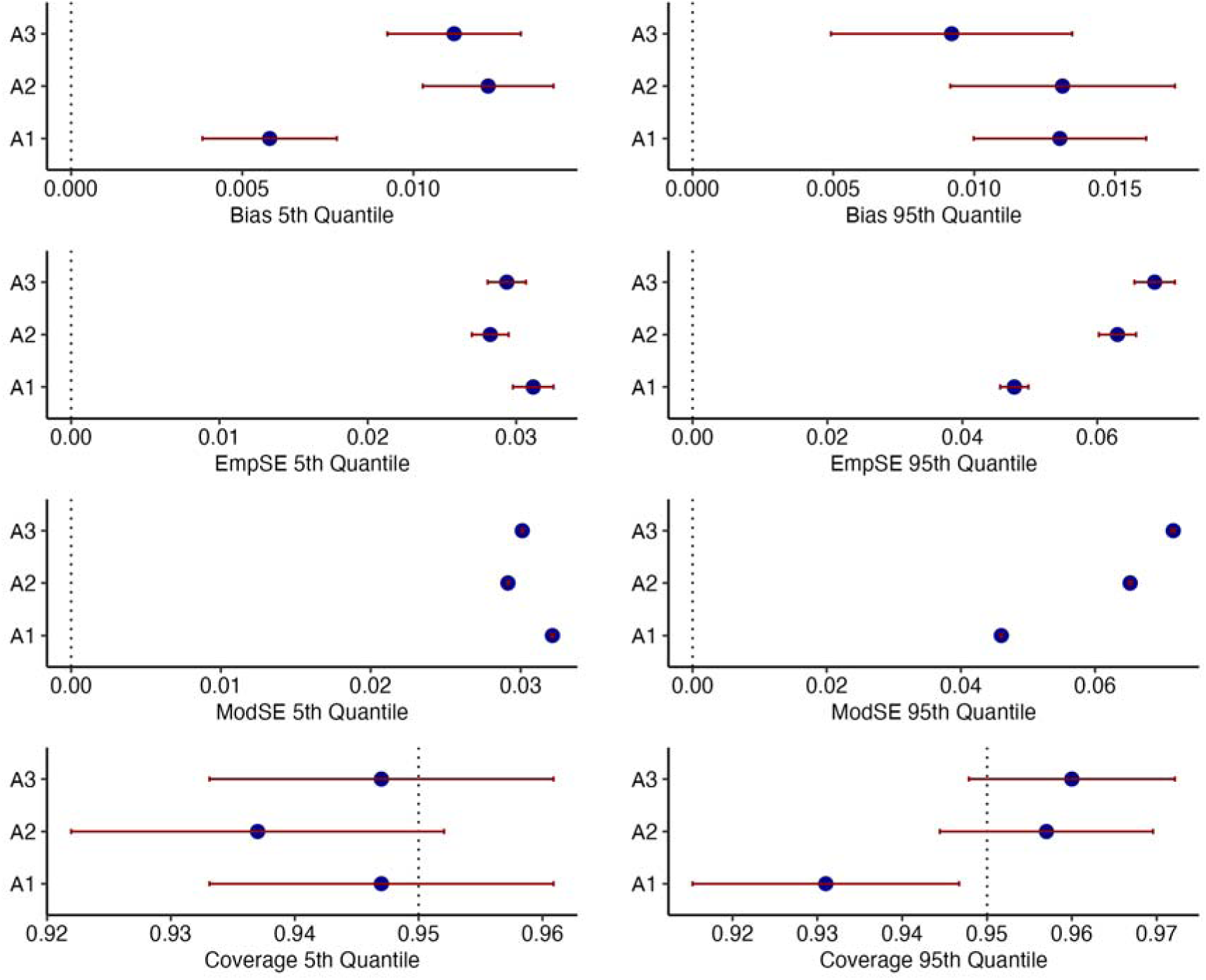
Performance measures and Monte Carlo 95% confidence intervals, with error bars, for the sensitivity of the main model (with 11 years of data, 24-hour lag and Poisson distribution) concerning lag and variable knots.

## 4. Discussion

This simulation study evaluates the performance of mf-DLNM in modelling the epidemiological associations between high-frequency exposure and low-frequency health outcomes. Hourly temperature data and daily respiratory mortality data from the West Midlands region were utilized to conduct this study, assessing the effectiveness of mf-DLNM for hourly-daily mixed frequency regression modelling. This study demonstrated that the mixed frequency mf-DLNM model performs well for hourly exposure and daily response data. Therefore, alongside available alternative methods, it offers a viable approach for modelling mixed-frequency regression.

The simulation results indicate that the mf-DLNM is a viable approach for mixed-frequency modelling in the absence of actual hourly health data. The main model, which utilized 11 years of daily respiratory mortality data and hourly temperature data, demonstrated lower bias and substantial coverage. The consistency of low bias across all scenarios suggests that this method could be effectively applied in situation where high frequency exposure data is available for low frequency health data. Among the six scenarios tested, the model incorporating seasonal variation (summer months with 11 years and 24-hour lag) showed suboptimal performance for the 95^th^ quantile of RR in terms of bias. However, coverage remained adequate. This performance discrepancy may be attributed to reduced data strength and the restriction of the dataset to a single season, rather than leveraging the full dataset for seasonal predictions. This could be improved with a large data size. However, small biases are acceptable due to substantial performance of measures, such as coverage.^24^

Previous studies have identified associations between elevated temperatures and adverse respiratory health effects, typically observed with a lag of 0 to 1 days.^27^ Additionally, a multicountry study found that the strongest associations between mortality and heatwaves were observed at 0-1 lag, diminishing by 3-4 lag.^28^ A study in northern U.S. cities (Boston, Philadelphia, Seattle) showed that the mortality risks peaked during late morning to evening hours (0900–2200 LST) at lag 0 (day of death) and lag 1 (previous day).^29^ For example, the study showed increased mortality risk at 0900 LST (lag 0) for temperatures at the 99th vs. 85th percentile in Philadelphia. A comparative analysis of daily and hourly temperature variability (TV) in relation to cardiorespiratory mortality, revealing that both TV_daily_ and TV_hourly_ had comparable impacts on mortality risk.^16^ However, due to the absence of high-resolution health data, most studies analysing the impact of hourly temperature do not incorporate complete temporal patterns into their models. Instead, they often rely on maximum temperatures, temperature ranges, or analyse each hour individually.^29–31^ As a result, these studies may not accurately capture the effect of hourly temperature variations on human health. Beyond temperature, other environmental variables have also been observed to exhibit associations with hourly changes in mortality and morbidity. A meta-analysis using 33 studies on intraday effects of ambient air pollution and temperature on cardiorespiratory morbidities demonstrated an association between total respiratory morbidity and hourly-lagged air pollution exposure.^32^ These findings underscore the importance of investigating hourly associations between environmental exposures and adverse health outcomes.

The validation of the mf-DLNM for hourly–daily data was undertaken with consideration of the broader implications of evaluating the health effects associated with hourly variations in environmental exposures. The development of mixed-frequency models is essential given the high temporal variability of environmental exposures and the limited availability of high-resolution health data. For instance, the wildfires in Los Angeles showed worsening of hourly air quality in the region.^33^ However, studies assessing the short-term risks of wildfire-related PM_2.5_ on cause-specific health outcome have typically been conducted at a daily temporal resolution, primarily due to the unavailability of hourly routine health data, even in events with known hourly changes.^34^ Recent studies still focus on assessing the daily association between air pollution^35^ and health outcomes, even when high temporal resolution hourly air pollution data is available.^36^ Furthermore, in comparison with high-income countries (HICs), low- and middle-income countries (LMICs) lack sufficient high-resolution health data.^37^ The most fine-grained temporal resolution for health data in LMICs often comes from daily or sub-daily data collection systems, therefore, using the mixed frequency model could provide high temporal hourly information on impact of exposures.

Furthermore, a broader objective of validating this model is to generate robust evidence on the relationship between hourly environmental exposures and health outcomes. This evidence could inform the development of targeted interventions to address hourly changes in temperature and air pollution, ultimately helping to mitigate the adverse health impacts associated with such variations. This could guide policymakers to build on interventions such as implementation of hyperlocal air quality and temperature tracking systems or GPS-based personal exposure monitoring to provide hourly risk notifications and warnings about high-risk zones.^38,39^ Furthermore, providing evidence on the importance of high-resolution information, highlights the need for accurately timed Electronic Health Records (EHR) data for both HIC and LMIC countries to have robust exposure-response modelling in environmental health research. In England, this issue primarily arises from structural and data governance limitations. Although the exact time of the event (e.g., arrival or admission) is typically recorded in the original dataset, the problem lies in how routine health data are processed and subsequently made available to researchers for linkage and analysis. Therefore, there is a need of structural and data improvement on how health dataset are provided to researchers.

The key strength of this study is a robust simulation analysis using an 11-year dataset and assessing various scenarios that consider different sample sizes, lag lengths, and specific seasons. However, it has certain methodological limitations. The key limitation of this study include the model constraint where the lag-response relationship for current modelling framework may not be able to capture the hourly lag individually, further advancement can be done using alternative modelling framework.^23^ In addition, one limitation is that the model was tested using only a single exposure–response pair, highlighting important challenges related to validation and generalizability. This raises concerns about whether the model’s performance metrics would remain consistent when applied to diverse exposure-response scenarios. Additionally, the model requires a sufficient data strength to effectively capture mixed-frequency associations, as observed through data-size-based scenario assessments. Furthermore, while this modelling framework specified knots and splines based on literature and sensitivity analyses, it may encounter difficulties when extended to more complex models involving multiple covariates. Although the current model utilized a large sample size, the model exhibited increased bias in estimating the RR of respiratory mortality for the 95th quantile of temperature. Large-scale simulations also require significant computational resources, which may pose a barrier to implementing complex models. Therefore, further evaluation addressing these shortcomings should be conducted in future research.

Furthermore, future studies should be conducted to assess the performance of a mixed frequency approach for other exposures such as air pollution (e.g., NO₂ and PM_2.5_) and other meteorological variables. Future investigations could explore alternative methodological frameworks for hourly and daily mixed-frequency data.^23^

## Conclusion

This simulation study validated the applicability of the mf-DLNM for high-resolution health risk estimation, even in the presence of data constraints. The study highlights the advantages of mf-DLNM as an alternative to traditional approaches that aggregate high-frequency exposure data, which often result in the loss of critical temporal information. By preserving the temporal information of environmental exposures, mf-DLNM provide more information on the possible impacts of short-term peaks of exposure on human health. These findings underscore the potential of mf-DLNM for advancing epidemiological research and public health risk assessment, particularly in scenarios where high-frequency exposure data are available, but corresponding health outcome data are available at lower temporal resolutions. Hence, a reliable high-resolution risk estimate improves healthcare preparedness and resource allocation during environmental episodic events.

## Data Availability

All data produced in the present study are available upon reasonable request to the authors.

